# Trends in the Incidence of Kaposi Sarcoma among Adults Attending HIV Care Facilities in East Africa and Latin America in the Treat-All Era

**DOI:** 10.64898/2026.06.26.26356374

**Authors:** Aggrey S. Semeere, Joshua Slone, Gustavo Amorim, Beverly Musick, Brenda Crabtree-Ramírez, Lameck Diero, Larissa Otero, Hilary Vansell Riley, Antony Ngeresa, Mark Nsumba, Haruna Ssemuwemba, Paul Enyel, Gertrude Nakigozi, Godloveness Rubega, Jerome Lwali, Guisela Salgado, Guilherme Calvet, Maria Fernanda Rodriguez, Ana Grana, Karen Juarez, Ran Tao, Stephany Duda, Constantin Yiannoutsos, Thomas Lumley, Jeffrey Martin, Pamela A. Shaw, Bryan E. Shepherd

**Author notes:** **Corresponding Author:** Aggrey S. Semeere, Infectious Diseases Insitute, P.O. Box 22418, Kampala, Uganda. **Funding:** R37 AI131771, U01 AI069923, U01 AI069911.

## Abstract

**Background:** In resource-rich regions, such as the U.S. and Europe, the incidence of Kaposi sarcoma (KS) amongst persons living with HIV (PWH) has dramatically declined with the advent of combination antiretroviral therapy (ART). In contrast, in low- and middle-income countries (LMICs), much less is known, particularly since the World Health Organization’s recommendation in late 2015 to use ART in all PWH. We take advantage of the coincident electronic clinical data capture at HIV care facilities to estimate the incidence of KS among PWH in care with ready access to ART, piloting a data validation approach to address errors in these routine clinic data.

**Methods:** We evaluated PWH enrolled from January 2010 to December 2019 in 13 HIV care clinics in 8 countries participating in the East Africa (EA-IeDEA) and Caribbean, Central and South America (CCASAnet) regions of the International Epidemiology Databases to Evaluate AIDS (IeDEA) consortium. Selected measurements were validated via chart review on a subset of PWH, and we estimated KS incidence in both unvalidated and validated data via generalized raking techniques.

**Results:** A total of 235,474 PWH from EA-IeDEA and 19,683 from CCASAnet gave rise to 719 and 103 incident cases of KS, respectively. A total of 824 eligible records were validated. ART use was substantially lower in EA-IeDEA than CCASAnet in 2010 but equalized by 2019. From 2010 to 2019, KS incidence decreased on average 21% per year (incidence rate ratio [IRR] 0.79; 95% CI 0.75-0.82) in EA-IeDEA but only 6% (IRR=0.94; 95% CI 0.83-1.06) in CCASAnet.

**Conclusions:** Among PWH attending HIV care facilities in East Africa, we observed a trend suggesting a reduction in KS incidence that paralleled increased Treat All era ART use in these clinics. In the Caribbean, Central and South America, there was hardly a change in the incidence, despite high-frequency ART use in the region as well.

## Introduction

Kaposi sarcoma (KS) is one of the most common cancers among people living with HIV (PWH). In resource-rich regions, such as the U.S. and Europe, the incidence of KS among PWH declined dramatically with the use of combination antiretroviral therapy (ART)(1–8). The U.S., for example, observed an estimated >50% decline in HIV-related KS incidence between 2000 and 2015 (1, 4). Earlier ART era data suggested initial declines in KS incidence and improved survival in low-and middle countries (LMIC)(9–14), although to a lesser magnitude than in resource-rich settings. Thus KS continues to occur among PWH with profound impact on morbidity and mortality (11, 15, 16). Since 2015, the implementation of the WHO Treat-All strategy means ART initiation occurs earlier (17), with more potent and durable antivirals (18), in an environment with improved KS screening and diagnosis (19–21). Within this context, incidence of KS among PWH warrants updating, particularly in populations with overlapping high prevelance of HIV and Kaposi Sarcoma Herpes Virus (KSHV) infections (22–25). Importantly, HIV-associated KS constitutes a majority of the new KS diagnosed (26), and yet KS incidence remains higher than pre-HIV epidemic estimates (15). Therefore after the wider ART roll-out, with PWH initiating ART irrespective of immunological status, it is unclear if KS incidence among PWH has even reduced further and if so to what extent.

Typically, such epidemiological assessments are done using population-based cancer registry data supported by underlying census information. While some cancer registries exist, most of them in sub-Saharan Africa have been deemed of insufficient quality for various reasons including not documenting HIV status (15, 27, 28). Representative HIV clinic databases containing routinely collected clinical data are an alternative that has been used to update KS incidence for PWH (10, 11, 13, 16) largely focusing on communities served by HIV clinics. Nonetheless data quality concerns also exist regarding the accurate capture of critical information like KS status and dates, within routine care, particularly in sites where KS diagnosis may be made in subspecialty clinics (29–31). We have previously demonstrated how to estimate incidence using such data (29, 32, 33). Regarding KS incidence estimation, an important aspect is that all KS diagnosis information should be captured correctly especially histopathological confirmation and dates. Errors in these critical data lead to inaccurate estimates, hence limiting the utility of routinely collected HIV care data.

In this paper we evaluate temporal trends in KS incidence after engagement in care among PWH in East Africa and Latin America during the decade that includes the initial implementation of the Treat-All strategy (2010–2019). We use data from two large and diverse HIV networks that rely on routinely collected HIV clinic data. Furthermore, given that these data arise from real-world settings with potential data quality issues (29–31), we employ recently developed statistical methods to sample records for data validation and to incorporate validated data to enhance the validity of our estimates (32, 33).

## Methods

### Cohort Description and Data Collection

We included data from 13 HIV clinical sites from 8 countries within two regions of the International Epidemiologic Databases to Evaluate AIDS (IeDEA) network (34). Participating sites from the Caribbean, Central and South America network for HIV Epidemiology (CCASAnet) (35) region of IeDEA were in Brazil, Chile, Honduras, Mexico, and Peru. Participating sites from East Africa (EA)-IeDEA (36) were in Kenya, Tanzania, and Uganda. Specific sites are included in the Supplemental Material.

Routinely collected clinical data from medical records were assembled at each site, de-identified, and sent to CCASAnet and EA-IeDEA data coordinating centers at Vanderbilt University Medical Center and Indiana University, respectively, for data harmonization and processing (37). Institutional ethics review boards from all participating sites and data coordinating centers approved the project (Vanderbilt IRB #060284 and #171353, and Indiana IRB #060478).

### Study Definitions and Outcomes

PWH were eligible if they were adults (≥18 years of age) at the time of study entry, defined as a participant’s first visit (i.e., enrollment date at HIV clinic) between January 1, 2010 and December 31, 2019 or January 1, 2010 for those with visits prior to 2010. PWH with no study visit from 2010-2019 were excluded. PWH diagnosed with KS before or within the 60 days after HIV clinic enrollment were also excluded.

KS diagnoses were based on histopathology and clinical diagnoses as recorded in patient records. Incident KS diagnoses were any new diagnoses made during the study period >60 days after enrollment in HIV care. Patient follow-up stopped at death, last visit if alive, or December 31, 2019 if death/last visit occurred after this date PWH were classified as lost to follow-up if their last visit was over a year prior to the first of December 31, 2019 or the site-specific database closing date, and they were not known to have died or transferred out of care (38); database closure was estimated as the date at which 95% of last visits occurred for the site.

### Data validation Processes

The Vanderbilt Data Coordinating Center randomly selected 1060 medical records for chart review using a two-wave stratified sampling strategy aimed to maximize precision of estimates by targeting the most informative records (39–41). Thirty-four strata were defined based on KS status and year of KS diagnosis (no KS, KS from 2010-2014, KS from 2015-2019), and study site (13 sites); because of sparse numbers, KS strata were collapsed across calendar time for 5 sites). Neyman allocation was used to determine how many to sample across strata; medical records with a KS diagnosis were over-sampled. Details of validation sampling procedures are provided elsewhere (42). Here we only report and incorporate validation data for records meeting inclusion criteria for our KS incidence analysis.

Data validation was performed by local auditors with knowledge of both clinical and data processes. All auditors attended a pre-audit training session. All sites received a specific list of IDs for which the auditors reviewed the primary data sources and checked for consistency with the analysis dataset, focusing on KS diagnosis and dates, ART initiation and dates, follow-up dates, demographics, and CD4 count measurements. Validation data were entered into REDCap. For several medical records, some variables were unable to be validated; for our purposes, we define a record as validated if KS diagnosis was validated. Failure to validate was mainly due to inability to locate physical medical records for review. Two records (both apparent discoveries of incident KS found during chart reviews) had a substantial impact on weighted analyses of validated data. These two records were further investigated, and it was determined that only one of the two actually had KS; the final validation dataset reflects these determinations.

### Statistical Analysis

We present two sets of analyses: those based on the original medical records (unvalidated data) and those based on weighted analyses of validated data. Estimates using validated data were weighted using calibrated inverse probability weights (IPW). IPW were calculated as one over the product of two estimated probabilities: the probability of being selected for validation and the probability of being validated given a record was selected for validation. The latter probability was incorporated to account for potential bias due to a selected chart not being able to be validated. The probability of being selected for validation was estimated using a logistic regression model including sampling strata as the covariate. The probability of being validated given selection for validation was estimated using logistic regression with last visit date, region, and death as covariates. To improve precision, IPW were calibrated with unvalidated ‘auxiliary’ data available for the entire study cohort using ‘generalized raking’ techniques. Generalized raking estimators are a special case of augmented IPW estimators (43). For Poisson regression analyses, the weights were calibrated using estimates of the influence function for each of the regression coefficients derived from fitting the models to the unvalidated cohort data. More details on generalized raking (44, 45) and its application to this study are presented elsewhere (42). For brevity, we refer to generalized raking estimators as ‘weighted’ estimators.

We provide incidence estimates stratified by region. We provide estimates of incidence by calender year of occurance using Poisson regression models treating year as 1) a categorical variable and 2) as a continuous variable expanded using natural splines with 3 knots. In adjusted analyses of incident KS, we fit Poisson models that included time-updated calendar year and controlled for sex at birth, region, time-fixed age at study entry, time-updated CD4 cell count (most recent prior measurement, square-root transformed and multiply imputed if missing), and a time-updated indicator of having started ART. These adjusted analyses, which are considered secondary, investigate whether trends seen in KS incidence can be explained by covariates, particularly ART use. Our primary incidence analyses based on the validated data excluded a single highly influential record (the record discovered to have KS during validation, described in *Data Validation Processes*). Sensitivity analyses included this highly inflential record.

Analyses were performed using `R`; specifically, validated generalized raking analysis used the `survey` package (46, 47). Analysis scripts are available at https://github.com/Joshua-Slone/KS-Clinical-manuscript.

## Results

### Study population

Our study included 255,157 PWH, with 19,683 from CCASAnet and 235,474 from EA-IeDEA (Figure 1). Of these records, 946 were selected for validation, 844 (89%) were validated, and 824 (87%) of the validated records remained eligible based on validated criteria, 91 from CCASAnet and 733 from EA-IeDEA (Supplementary Table 1). Based on chart reviews, KS diagnosis was in error in 0.6% of eligible validated records (4 false positives and 1 false negative; details in Supplementary Table 2), date of KS diagnosis was in error in 4.2% of validated incident KS diagnoses, date of ART initiation was in error in 7.8% of validated records (17 false negatives and 47 different dates), sex was in error in 0.2% of validated records (2 records changed from male to female sex after validation), age was in error in 7.4% of validated records, and CD4 count at beginning of follow-up was in error in 4.1% of validated records.

**Figure 1.**
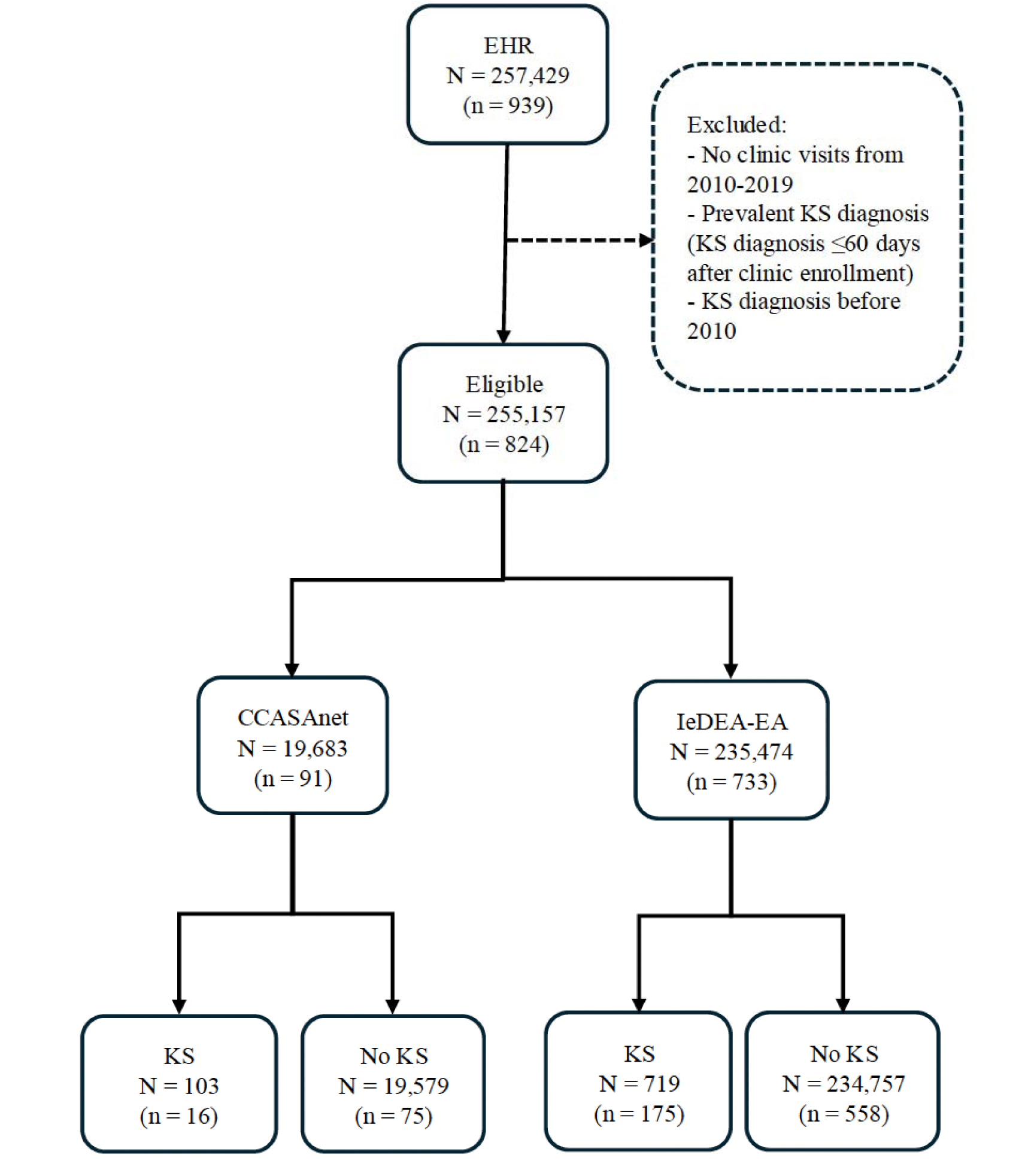
Inclusion diagram for study. **Footnote: ‘**N’ refers to unvalidated cohort while ‘n’ in the parentheses refers to unweighted validated records for each outcome.

Characteristics of PWH included in this study are in Table 1, which includes unvalidated data for the entire study population, and weighted estimates for the study population based on the validated cohorts. The EA-IeDEA cohort was approximately 34% male, whereas the CCASAnet cohort was predominantly male (78%). The proportions of those who were on ART at time-zero (EA-IeDEA 37.3%, CCASAnet 36.5%) and during follow-up (weighted estimates, 79% in EA-IeDEA and 81% in CCASAnet) were similar between cohorts. Median follow-up was a little over 3 years with a total of approximately 1 million person years of follow-up. Loss to follow-up was 42% for EA-IeDEA and 33% for CCASAnet.

**Table 1:** Characteristics of the A) EA-IeDEA and B) CCASAnet study populations stratified by incident KS; estimates are presented based on unvalidated data in the entire cohort and based on the validated data weighted to represent the larger cohort.

**A. EA-IeDEA**
| Variable | Entire non-validated cohort |  | Weighted validated cohort |  |
| --- | --- | --- | --- | --- |
|  | Incident KS | No Incident KS | Incident KS | No Incident KS |
|  | (N=719) | (N=234,755) | (n=175) | (n=558) |
| Male sex<br>(vs. female sex) | 53% | 34% | 48% | 34% |
| Age at start of follow-up | 34.9 (29.7, 41.3) | 34.9 (28.3, 42.4) | 34.5 (28.1, 40.2) | 36.0 (29.2, 42) |
| Year follow-up began | 2010 ('10, '13) | 2012 ('10, '14) | 2010 ('10, '12) | 2012 ('10, '15) |
| 2010-2014 | 87% | 75% | 89% | 74% |
| 2015-2019 | 13% | 25% | 11% | 26% |
| CD4 count at start of follow-up | 254 (92, 432) | 411 (225, 534) | 235 (108, 357) | 394 (238, 538) |
| ART at start of follow-up<br>(vs. no ART at start) | 38% | 41% | 37% | 39% |
| Ever on ART<br>(vs. never on ART) | 79% | 78% | 79% | 85% |
| Follow-up time (years) | 0.6 (0.3, 1.9) | 3.1 (0.6, 7.3) | 0.7 (0.3, 1.7) | 3.5 (0.7, 7.7) |

**B. CCASAnet**
| Variable | Entire non-validated cohort |  | Weighted validated cohort |  |
| --- | --- | --- | --- | --- |
|  | Incident KS | No Incident KS | Incident KS | No Incident KS |
|  | (N=103) | (N=19,580) | (n=16) | (n=75) |
| Male sex<br>(vs. female sex) | 96% | 77% | 96% | 74% |
| Age at start of follow-up | 34.1 (28.2, 39.3) | 34.8 (27.9, 43.6) | 28 (24.9, 51.9) | 34.6 (28.6, 43.6) |
| Year follow-up began | 2011 ('10, '14) | 2012 ('10, '16) | 2011 ('10, '13) | 2012 ('10, '15) |
| 2010-2014 | 81% | 65% | 81% | 75% |
| 2015-2019 | 19% | 35% | 19% | 25% |
| CD4 count at start of follow-up | 134 (44, 349) | 356 (180, 538) | 90 (18, 505) | 453 (369, 802) |
| ART at start of follow-up<br>(vs. no ART at start) | 36% | 46% | 37% | 45% |
| Ever on ART<br>(vs. never on ART) | 83% | 87% | 81% | 91% |
| Follow-up time (years) | 0.9 (0.3, 3.1) | 4.3 (1.3, 7.9) | 1.8 (0.6, 3.7) | 5.1 (2.7, 7.8) |

Binary variables include percentages. Numeric variables include medians (with 25th and 75th percentiles).

### KS Incidence and Trends

In EA-IeDEA we observed 719 incident KS diagnoses while in CCASAnet we observed 103. These new diagnoses represent an average incidence of 79 (95% CI: 73-85) and 115 (95% CI: 95-139) KS cases per 100,000 person-years in EA-IeDEA and CCASAnet, respectively, during the study period. In weighted analyses incorporating validated data we obtained similar results: 76 (95% CI 72-79) and 101 (95% CI: 81-127) KS cases per 100,000 person-years in EA-IeDEA and CCASAnet, respectively.

Estimated incidences by year are presented in Figure 2, with the top and bottom panels corresponding to the unvalidated and weighted validated analyses, respectively. In EA-IeDEA, there was a downward trend during the first half of the decade and then a stable low, but non-zero, incidence of KS in the latter half. Using both the unvalidated and validated data, the incidence of KS in EA-IeDEA was estimated to decrease on average from 2010 to 2019 by about 21% per year (IRR=0.79; (95% CI 0.76-0.81) and IRR=0.79 (95% CI 0.75-0.82) respectively; Table 2). In contrast, trends in the incidence of KS in CCASAnet were much less pronounced over the decade: IRR=0.93 per year (95% CI 0.86-1.00) in both unvalidated and with validated data (0.94 (95% CI 0.83-1.06)).

**Figure 2:**
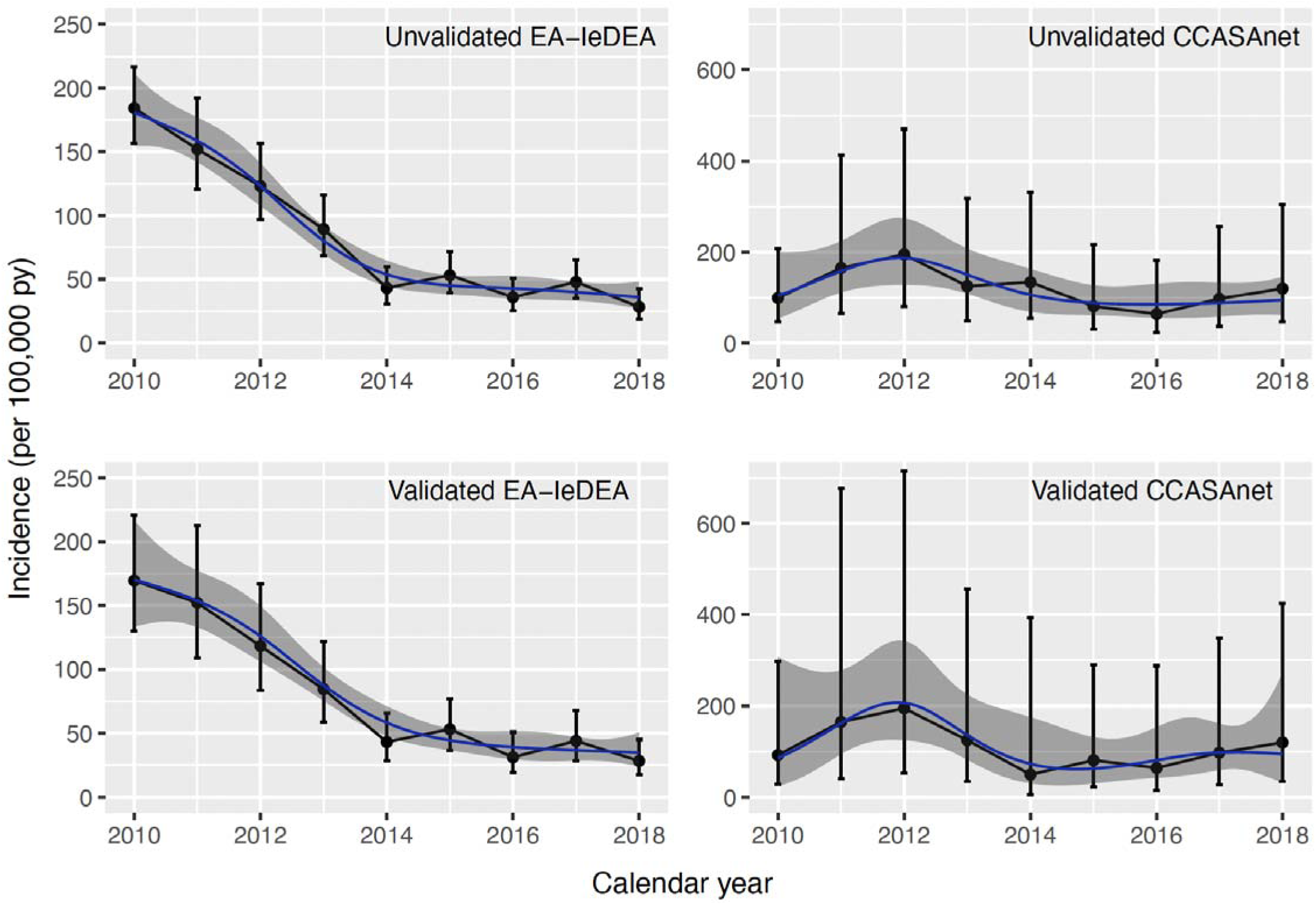
Estimated KS incidence rate trends over time in EA-IeDEA (left column) and CCASAnet (right column) using the unvalidated phase-1 data (first row) and the weighted validation data (second row). Estimates based on a natural spline and 95% CI are included. The EA-IeDEA weighted validation panel excludes one highly influential record; Supplementary Figure 1 shows the curve with this record included. Note that different y-axis scales were used for the two regions to make trends more apparent.

**Table 2.** Incidence, Incidence Rate Ratios (IRRs), and 95% confidence intervals for Kaposi arcoma in East Africa and Latin America based on the unvalidated medical record data and generalized raking estimates using validated data. The EA-IeDEA weighted validated data column excludes one highly influential record; Supplementary Table 3 shows estimates with this record included.

|  | EA-IeDEA |  |  |  | CCASAnet |  |  |  |
| --- | --- | --- | --- | --- | --- | --- | --- | --- |
|  | Unvalidated Data |  | Validated Data |  | Unvalidated Data |  | Validated Data |  |
| Incidence (per 100,000 py) |  |  |  |  |  |  |  |  |
| Overall | 79 | (73-85) | 76 | (72-79) | 115 | (95-139) | 101 | (81-127) |
| Before ART initiation | 96 | (82-113) | 96 | (94-98) | 268 | (169-426) | 163 | (38-697) |
| After ART initiation | 75 | (69-82) | 72 | (68-76) | 102 | (83-127) | 84 | (45-156) |
| ≤ 90 days | 451 | (380-535) | 415 | (354-486) | 815 | (514-1294) | 443 | (155-1266) |
| > 90 days | 60 | (55-66) | 58 | (54-62) | 83 | (65-105) | 82 | (77-87) |
| Calendar Year IRR (per year) |  |  |  |  |  |  |  |  |
| Unadjusted | 0.79 | (0.76-0.81) | 0.79 | (0.75-0.82) | 0.93 | (0.86-1.00) | 0.94 | (0.83-1.06) |
| Adjusted* | 0.80 | (0.77-0.84) | 0.82 | (0.78-0.86) | 0.95 | (0.88-1.03) | 0.92 | (0.74-1.16) |

\* Adjusted for sex, age, ART status, and CD4 cell count.

### ART use and incident KS

During the study period, the proportion on ART increased in both cohorts (Figure 3). At time of database closure over 95% of PWH were on ART despite starting lower in EA-IeDEA (<70%) than CCASAnet (>80%). Among PWH on ART in EA-IeDEA, KS incidence was lower (72 per 100,000 person-year; 95% CI: 68-76 with validated data – results were similar with unvalidated data), compared to incidence before ART initiation (96, 95% CI: 94-98 with validated data). However, during the first 90 days after ART initiation, the incidence of KS was high (415 per 100,000 person-years, 95% CI: 354-486 with validated data), and then dropped substantially therafter (58 per 100,000 person-years, 95% CI: 54-62 with validated data). A similar trend was observed in CCASAnet (Table 2).

**Figure 3:**
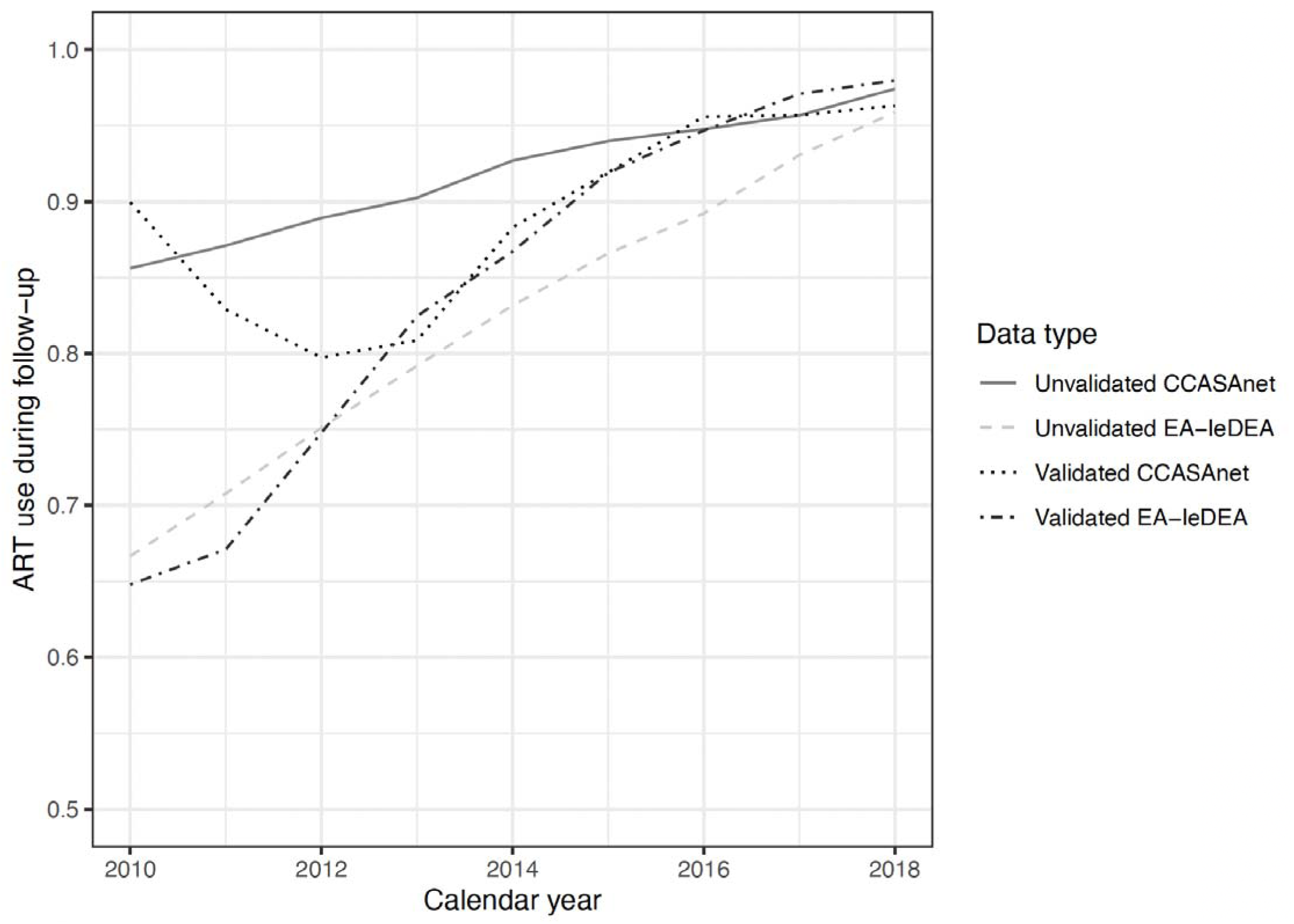
**Percentage of follow-up time each year on ART for both cohorts based on the unvalidated (phase-1) data and the validated data.**

In analyses that adjusted for variables potentially associated with incident KS including sex, age, ART status, and CD4 count, calendar trends for KS were similar in both validated and unvalidated data in both regions. In EA-IeDEA we estimated an adjusted Incident Rate Ratio of 0.80 per year, (95% CI: 0.77-0.84) in unvalidated data, and 0.82 per year (95% CI 0.78-0.86) with validated data. Similarly in CCASAnet we had adjusted Incident Rate Ratio of 0.95 per year (95% CI 0.88-1.03) with unvalidated data, and 0.92 per year (95% CI 0.74-1.16) with validated data.

As mentioned in Methods, one record belonging to a male in EA-IeDEA discovered to have KS, had an unrealistically large impact on weighted validated incidence estimates and was removed from primary incidence analyses. In sensitivity analyses, we re-computed estimates including this record (Supplementary Figure 1 and Supplementary Table 3). The overall incidence estimate in EA-IeDEA with this record included was higher, 100 per 100,000 person years, and because this person had started ART for >90 days at the time of his KS diagnosis in 2018, incidence estimates on ART, >90 days after ART initation, and in 2018 similarly increased. Because of the increased estimated incidence in 2018, when summarizing the curve with a line, the overall trend became essentially flat (unadjusted IRR of 0.97 per year; 95% CI 0.72-1.32).

## Discussion

Using routine clinic data, we update estimates of KS incidence for PWH in care in East Africa and Latin America between 2010 and 2019 using IeDEA-affiliated clinics. With extensive data validation and employing a novel generalized raking analysis approach in error prone data, we addressed data quality challenges in routinely collected clinic data for this at-risk population over a 10-year period. Overall, we observed higher KS incidence among PWH in Latin American clinics compared to East African irrespective of ART use. KS incidence reduced with increasing access to ART in East Africa with little evidence of reduction in Latin America over this calender period. Incidence in East Africa leveled off towards the end of the study period but was not down to zero. Below, we provide some contextualization of these findings in light of related research and other critical considerations.

Between 2010 and 2019 in East Africa, with validated data we estimated KS incidence as 76 per 100,000 person-years among PWH in care. In East Africa, this KS incidence estimate was much lower than the 321 per 100,000 person-years estimated between 2007 to 2012 (15), suggesting an overall decline between the two periods. Similarly, incidence was still lower than the rate for the period 2004-2010 observed previously in a similar setting (164 per 100,000 person-years), further suggesting possible reduced KS risk among adult PWH in care in East Africa (13). Specifically, after ART initiation, we estimated a weighted incidence of 72 per 100,000 person-years (Table 2) which is also lower than previous estimates that ranged between 138 to 340 per 100,000 person years, among PWH in LMICs prior to the current calender period (15).

In comparision, overall the weighted KS incidence among PWH in the Latin America clinics were higher. KS incidence was 101 per 100,000 person years, and 84 per 100,000 person years after ART initiation using the validated data. This difference in incidence could be largely explained by the unique differences in the underlying populations of PWH between the regions. In Latin American, PWH in care within the CCASAnet cohort are predominantly male (78%) populations while the EA-IeDEA population is predominantly female (65%). First, males in general are at higher risk of KS compared to females, irrespective of HIV status and this may in part might explain the higher incidence in the CCASAnet population (48). Further, male PWH in Latin America are more likely men-who-have-sex-with-men (MSM) with higher risk of Kaposi sarcoma associated herpesvirus (KSHV) infection and hence a higher likelihood of KS (48–50).

Contextualising our KS incidence estimate from Latin America is limited by the relatively fewer previous KS incidence estimates among PWH to compare with from the region. An earlier CCASAnet study that covered the years 2005 to 2017 estimated KS incidence was between 300-500 per 100,000 person-years among PWH (11). These estimates were higher than our 101 per 100,000 person-years estimated in Latin America. In this prior study, incident KS included diagnoses at clinic entry accounting for 22% of their diagnoses. Our current study excluded these cases; this may explain some of the discrepancy between estimates. In addition, the earlier CCASAnet study had two additional sites (one in Argentina and another in Brazil). The stability in KS incidence seen from 2005 to 2017 in the earlier paper is consistent with that observed in the current paper (11). The relatively stable KS incidence could in part be explained by the fact that over this study period, ART use in CCASAnet increased only slightly (Figure 3).

Conversly, in East Africa, incidence of KS in the treat-all era continued to decline among PWH in care. Across the study period, we observed an average 0.79 times lower incidence of KS per year. More specifically, there was a larger magnitude of decline observed between 2010 and 2014, and this leveled-off from 2015-2019, ranging from approximately 25-50 per 100,000 person years. This reduction was in tandem with increasing ART use that was estimated at 65% in 2010 to over 95% in 2019. Importantly, KS incidence is not down to zero despite the increased access to ART, suggesting that even with Treat-All, KS continues to occur among PWH mostly on ART.

KS incidence was highest in the first 90 days after ART start, suggesting that new KS could be due to the paradoxical or unmasking KS Immune Reconstitution Inflammatory Syndrome (IRIS) in response to ART initiation (51, 52). KS IRIS is likely since early KS is symptomatically subtle and hence clinically mild or inconspicuous to diagnose. In such cases KS might only get diagnosed after ART initiation when the aroused immune system makes it overt and symptomatic. Therefore the symptomatology after ART initiation precipitates diagnosis, yet the KS occurred prior to ART. Without detailed clinical and laboratory assessments, with routine clinic data we cannot reliably determine when exactly the KS among such invidivudals started in relation to ART.

Interestingly, the observed calendar year trends in KS incidence remained similar in secondary analyses that adjusted for demographics, CD4 count, and time-varying ART use (Table 2). Taken at face value, this suggests that the trends cannot be explained by changes in individual ART use. This result is surprising, and should not be over-interpreted. Potential explanations could include poor compliance to antiretrovirals (data on adherence or HIV RNA after ART initiation were limited) or trends in earlier enrollment in care (CD4 count data were somewhat sporadically collected and may not have sufficiently captured disease stage). These explanations are hypothetical and warrant further investigation.

Our study has additional strengths and limitations, that should be appreciated while interpreting our findings. We included data from over 250,000 PWH from diverse LMIC clinic settings. We performed extensive chart reviews to validate data on over 800 of these people, and then we incorporated the chart review data into analyses with the larger dataset using modern statistical methods to reduce bias due to the potentially error-prone nature of the routinely collected data. This extensive data quality work is novel and gives us greater confidence in our estimates. However, despite extensive chart reviews, data errors remain a possibility. In addition, in our primary analyses we removed a single discovered incident KS case in EA-IeDEA because of its extreme influence on study results; suggesting that our novel statistical methods using validation data to enhance estimates are still challenged by rare outcomes and rare errors. We observed incidence differences between Latin American and East African sites. While there could be better KS diagnostic procedures in Latin America, there are also, non-ignorable regional population differences among PWH which could explain the differences in incidence, as discussed above.

In both Latin America and East Africa, KS remains an important comorbidity among PWH. Overall, from 2010-2019 we observed a declining trend in incidence among PWH in care in East Africa and little to no change in Latin America. With over 95% ART access, incidence was still higher among men and there is a slower to no change in KS incidence in recent years suggesting a need to consider why and whether alternative strategies are needed to further reduce occurance.

## Data Availability

The data for this study arose from patient care and is, thus, protected and not available publicly.

## Supplementary Material

Participating sites in Latin America were Instituto Nacional de Infectologia - Evandro Chagas, Fundação Oswaldo Cruz, Rio de Janeiro, Brazil (Brazil-INI); Fundacion Arriaran, Santiago-Chile (Chile-FA); Instituto Hondureño de Seguridad Social and Hospital Escuela, Tegucigalpa, Honduras (Honduras-IHSS/HE); Instituto Nacional de Ciencias Médicas y Nutrición Salvador Zubirán, Mexico City, Mexico (Mexico-INCMNSZ); and Instituto de Medicina Tropical Alexander von Humboldt, Lima, Perú (Peru-IMTAvH). Participating sites in East Africa were Academic Model Providing Access to Healthcare (AMPATH), Eldoret, Kenya; Family AIDS Care & Education Services (FACES), Kisumu, Kenya; Infectious Disease Institute (IDI), Kampala, Uganda; Masaka Regional Hospital HIV Care Clinic, Masaka City, Uganda; Mbarara University ISS Clinic, Mbarara, Uganda; Morogoro Regional Hospital CTC, Morogoro Region, Tanzania; Rakai Health Sciences Program, Kyotera District, Uganda; and Tumbi Regional Referral Hospital CTC, Kibaha Town, Pwani Region, Tanzania.

---

**Supplementary Table 1.** Characteristics of the 946 records selected for validation, classified by whether the records were subsequently validated.

| Variable | All selected records for incidence analyses |  |
| --- | --- | --- |
|  | Records selected<br>but not validated<br>(*n*=102) | Records selected<br>and validated<br>(*n*=844) |
| EA-IeDEA<br>(vs. CCASAnet) | 96% | 89% |
| Male sex<br>(vs. female sex) | 46% | 42% |
| Age at start of follow-up | 33.7 (27.6, 44.0) | 35.2 (28.7, 42.0) |
| Year follow-up began | 2011 ('10, '13) | 2011 ('10, '14) |
| 2010-2014 | 84% | 75% |
| 2015-2019 | 16% | 25% |
| CD4 count at start of follow-up | 310 (104, 476) | 341 (164, 508) |
| ART at start of follow-up<br>(vs. no ART at start) | 39% | 39% |
| Ever on ART<br>(vs. never on ART) | 68% | 84% |
| Follow-up time (years) | 0.9 (0.2, 2.0) | 2.5 (0.5, 6.6) |

**Footnote:** Of the 844 records that were validated, 824 were eligible based on the validated criteria. The 20 ineligible records after validation were all KS cases that became prevalent cases (and thus ineligible) after validation due to updated dates of KS diagnosis and/or updated dates of enrollment in care.

**Supplementary Table 2.**
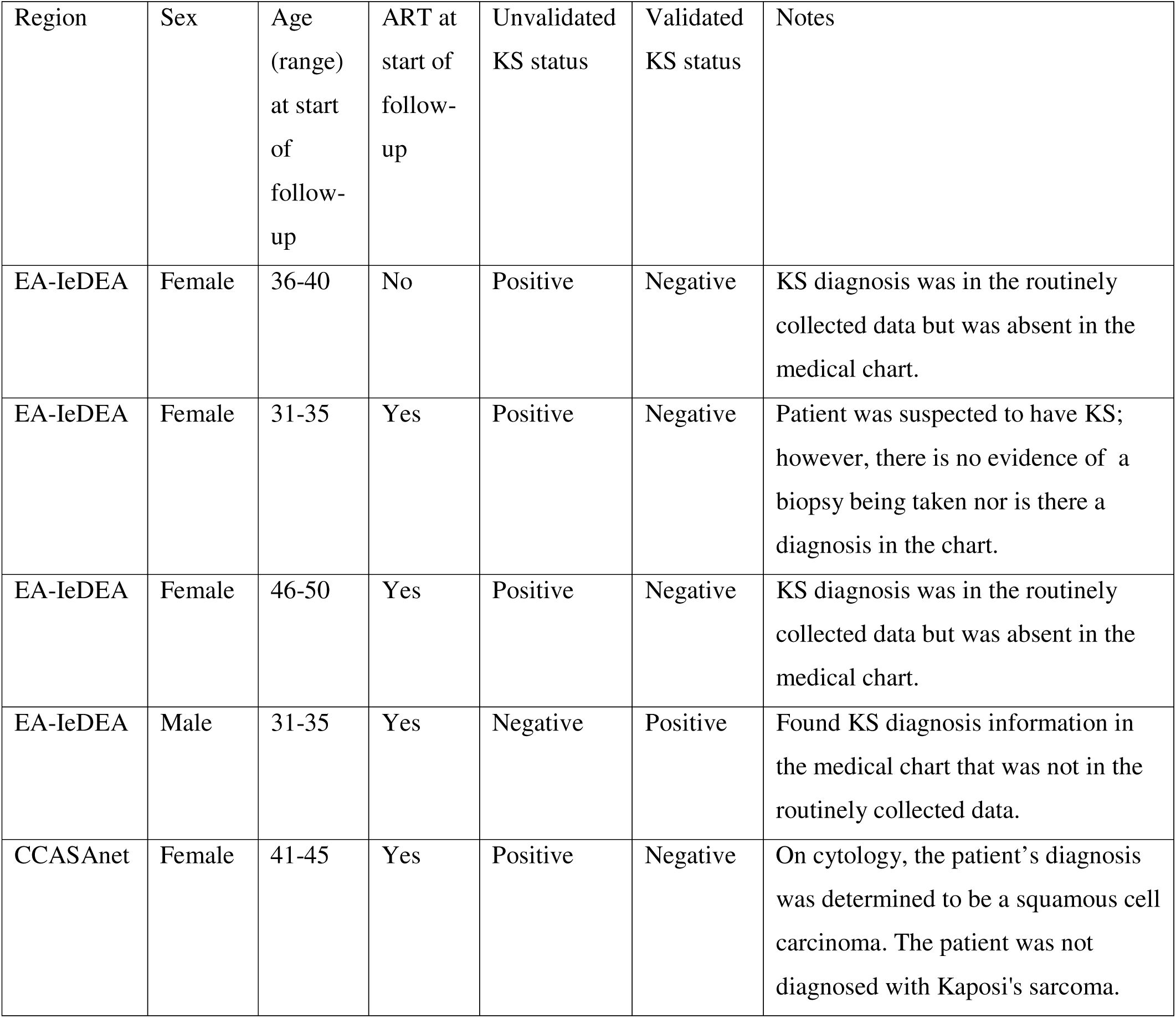
Information on records that went from KS diagnosed case in the unvalidated data to no diagnosis of KS in the validation, or no diagnosis of KS in the unvalidated data to KS diagnosed case in the validation.

**Supplementary Table 3.**
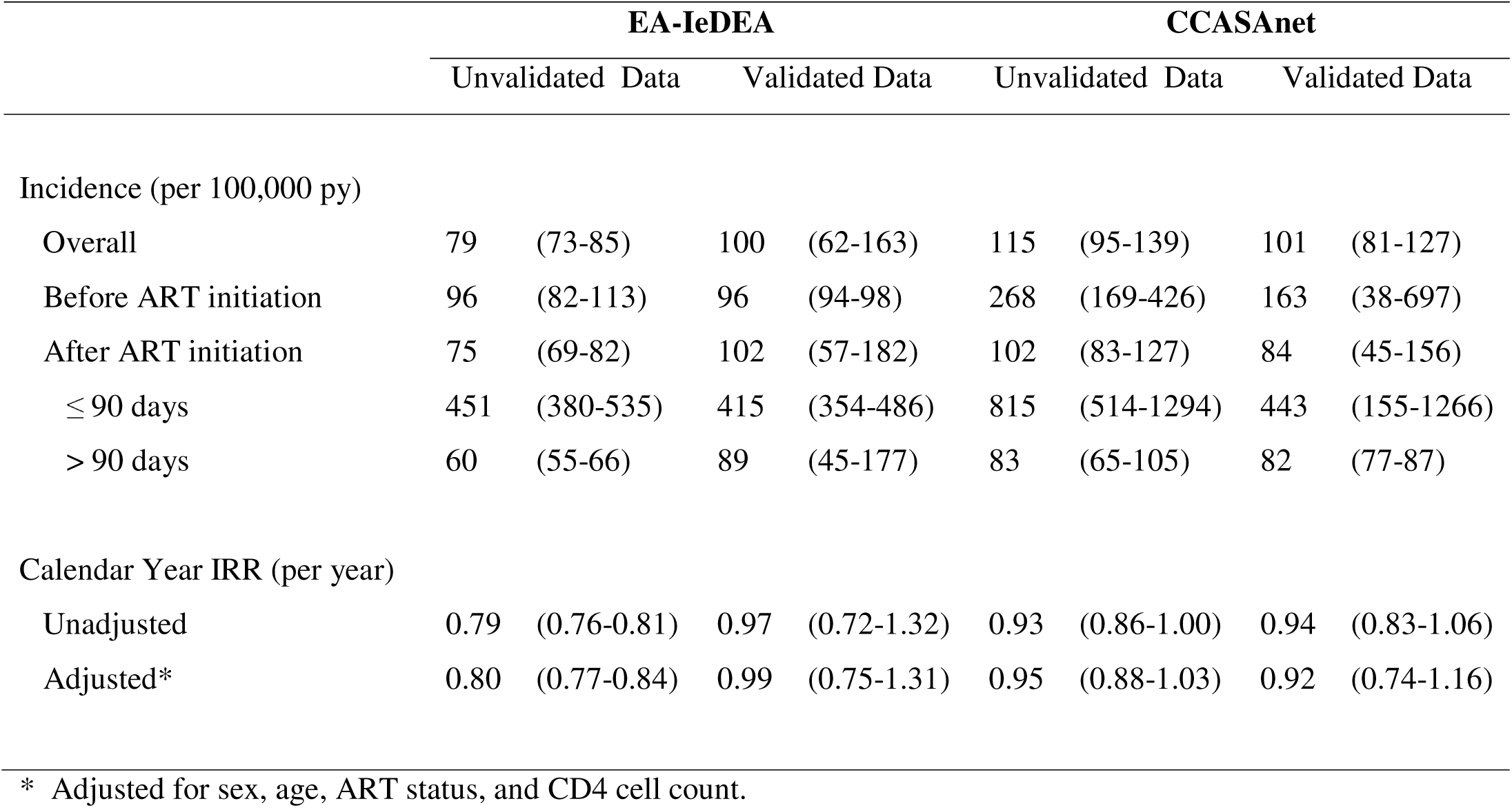
Incidence, Incidence Rate Ratios (IRRs), and 95% confidence intervals for Kaposi arcoma in East Africa and Latin America based on the unvalidated medical record data and generalized raking estimates using validated data. The EA-IeDEA weighted validated data column **includes** one highly influential record; Table 2 in the main manuscript shows estimates with this record excluded.

**Supplementary Figure 1:**
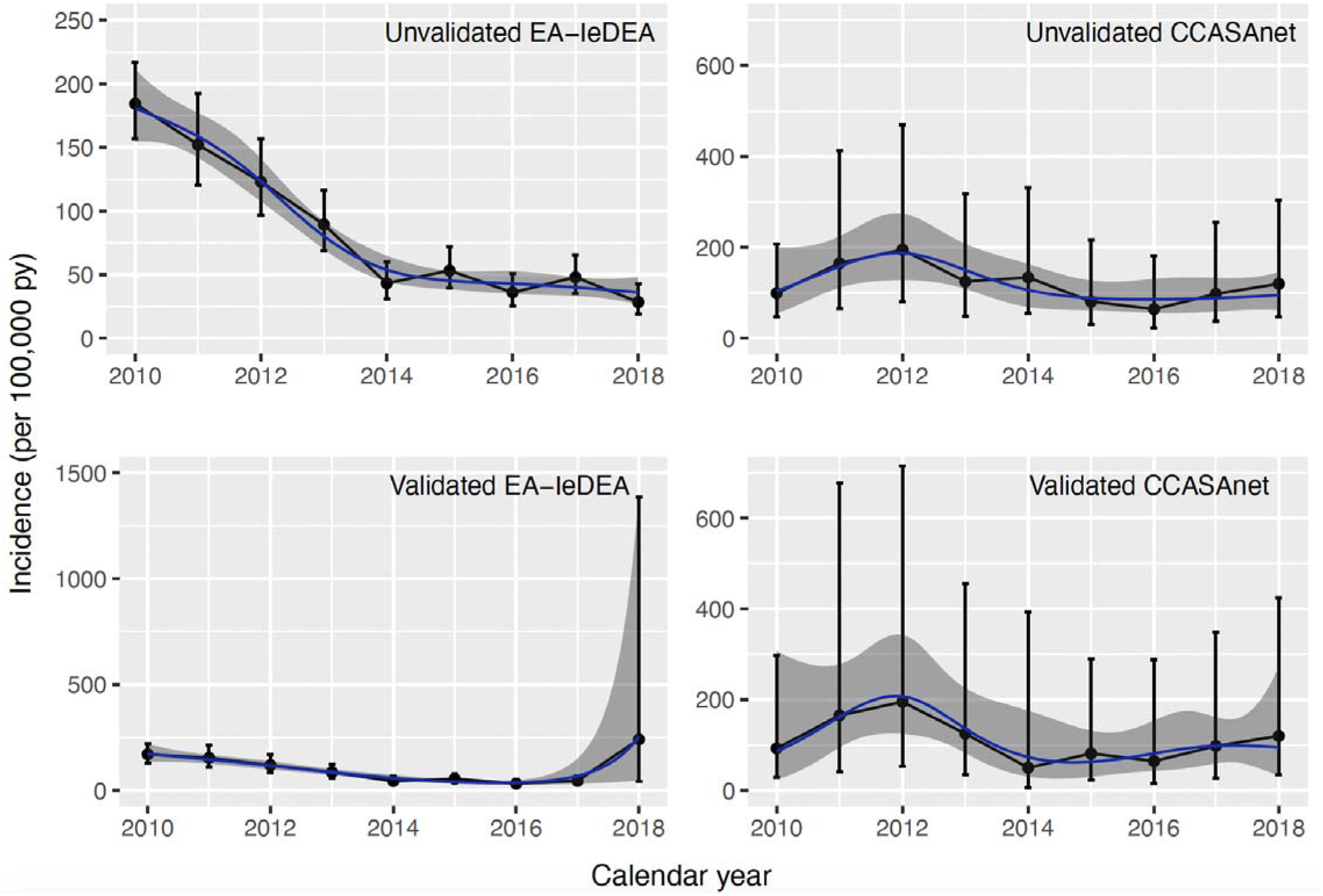
Estimated KS incidence rate trends over time in EA-IeDEA (left column) and CCASAnet (right column) using the unvalidated phase-1 data (first row) and the weighted validation data (second row). Estimates based on a natural spline and 95% CI are included. The EA-IeDEA weighted validation panel includes one highly influential record; Figure 2 in the manuscript shows the curve with this record excluded

